# Clinical course and outcomes of antibody-mediated rejection after heart transplant in the contemporary era

**DOI:** 10.64898/2026.05.19.26353576

**Authors:** Bin Q Yang, Mahmoud Elesawy, Sofia Laux, Elena Deych, Amanda Fernandes, Vikram Pattanayak, Kristen E Wong, Lana Tsao, Daniel A Zlotoff, Antonia Kreso, Joel D Schilling, Gregory D Lewis

## Abstract

**Background:** Antibody-mediated rejection (AMR) after heart transplant (HT) is associated with increased risk of mortality and graft loss. Contemporary studies delineating AMR presentation, management, and response to treatment are lacking, especially for patients who do not have typical immunohistological evidence of rejection (biopsy-negative, BN-AMR). In this study, we sought to describe the prevalence and clinical course of BN-AMR compared to biopsy-positive (BP-AMR) patients in a multicenter HT population.

**Methods:** We conducted a retrospective analysis of all adult HT recipients at 2 academic medical centers. AMR was further divided into BP-AMR and BN-AMR, depending on their endomyocardial biopsy findings. The primary outcome was death and secondary outcome was a composite of death, retransplant, and new International Society of Heart and Lung Transplant grade 2 or 3 coronary artery vasculopathy.

**Results:** A total of 742 patients were included in this study. We found that AMR occurred in 10% of HT recipients and was associated with worse overall survival compared to those with only cellular rejection or no rejection. BN-AMR accounted for 33% of AMR cases. Compared to BP-AMR, BN-AMR was diagnosed later, less aggressively treated, and associated with high morbidity and mortality. The long-term outcomes between BP-AMR and BN-AMR were similarly poor, with 5-year mortality approaching 50% after diagnosis.

**Conclusions:** AMR after HT is associated with poor clinical outcomes and BN-AMR is common. Future studies should focus on incorporating novel tools for earlier detection of AMR and investigating AMR sub-phenotypes and optimal modes of treatment.

## Introduction

Antibody-mediated rejection (AMR) is associated with increased risk of mortality and graft loss after heart transplant (HT).^1-8^ AMR pathology is characterized by antibody deposition on capillary endothelium, infiltration of mononuclear cells, and complement activation. Historically, AMR diagnosis is made by histological examination of endomyocardial biopsies (EMBx), in conjunction with immunohistochemistry and detection of circulating donor-specific antibodies (DSAs).^9-11^ However, AMR is increasingly recognized as a spectrum of disease ranging from acute allograft dysfunction to chronic coronary artery vasculopathy (CAV). Moreover, AMR can occur in absence of complement activation or DSAs.^12-15^ As such, the true prevalence of AMR is likely underestimated, with studies showing that AMR may be implicated in 30-50% of late-failing allografts.^16,17^

Advances in immunosuppression and surveillance strategies that incorporate routine screening for DSAs and use of donor-derived cell-free DNA (dd-cfDNA) have led to earlier detection of pathology in the HT population.^10,18-23^ However, patients with AMR continue to have poor clinical outcomes, although contemporary studies detailing management and clinical trajectories of these patients are lacking. In addition, treatment of AMR is highly variable across HT centers. In particular, HT recipients with clinical heart failure symptoms or DSAs with a negative biopsy are especially challenging to manage.^10,11^ Likewise, in a survey of HT providers, 20-50% of respondents reported not treating histologically diagnosed AMR without graft dysfunction.^24^ Thus, the clinical and laboratory findings that should prompt treatment remain controversial. Although there is no consensus treatment protocol, the management of AMR typically focuses on antibody removal via plasmapheresis/exchange (PLEX), antibody neutralization via intravenous immunoglobulin (IVIG), and biologic therapeutics targeting various pathways involved in antibody production and immune response.^9,25-29^

In this study, we sought to describe the presentation, treatment, and clinical course of AMR in a multicenter HT cohort. We further stratified AMR by the presence (biopsy-positive, BP-AMR) or absence (biopsy-negative, BN-AMR) of immunohistological evidence of rejection. We hypothesized that BN-AMR patients would be undertreated and have worse clinical outcomes than BP-AMR.

## Methods

### Patient population

All adult patients who underwent *de novo* HT at Barnes-Jewish Hospital/Washington University in St. Louis between 1/2010 and 8/2020 and Massachusetts General Hospital/Harvard Medical School between 1/2016 and 9/2023 were included in this study. All data were obtained from institutional HT databases,The IRBs at both centers approved this study and this study is in compliance with the ISHLT ethics statement.

### Post-HT care

All HT recipients during the study period were initiated on 3-drug immunosuppression with tacrolimus, mycophenolate mofetil, and steroids after transplant surgery and were monitored for rejection with routine surveillance EMBx that occurred weekly at first but decreased in frequency thereafter (**Supplemental table 1**). Induction therapy was not routinely used, although was employed for highly sensitized patients or those deemed to be at high immunological risk. Per protocol, steroids were gradually weaned over the first 6 months post-transplant although immunosuppression adjustments were made at the discretion of the treating physician. Standard prophylaxis against pneumocystis/toxoplasmosis, fungal infections, and cytomegalovirus were employed for the first 3 months post-HT.

**Table 1.** Baseline characteristics of HT recipients. ACR: acute cellular rejection, AMR: antibody-mediated rejection, high risk cytomegalovirus (CMV) mismatch: donor CMV IgG+ and recipient CMV IgG-, PRA: panel reactive antibodies.

|  | No rejection<br>(N=480) | ACR<br>(N=189) | AMR<br>(N=73) | P-value |
| --- | --- | --- | --- | --- |
| **Age, years (std dev)** | 56 (11) | 54 (12) | 48 (13) | <0.01 |
| **Male sex, % (n)** | 73 (351) | 69 (130) | 62 (45) | 0.10 |
| **Race, % (n)** |  |  |  | <0.01 |
| White | 78 (376) | 80 (151) | 53 (39) |  |
| Black | 17 (79) | 14 (26) | 44 (32) |  |
| Other | 5 (25) | 6 (12) | 3 (2) |  |
| **Body mass index, kg/m<sup>2</sup> (std dev)** | 28.1 (5.0) | 27.7 (4.5) | 29.2 (4.5) | 0.15 |
| **Heart failure etiology, %ischemic (n)** | 35 (170) | 32 (61) | 12 (9) | <0.01 |
| **Hypertension, %yes (n)** | 38 (184) | 41 (77) | 36 (26) | 0.68 |
| **Diabetes, %yes (n)** | 32 (154) | 30 (56) | 28 (20) | 0.64 |
| **Chronic kidney disease, %yes (n)** | 41 (199) | 28 (53) | 42 (31) | <0.01 |
| **High risk CMV mismatch, %yes (n)** | 29 (141) | 28 (53) | 18 (13) | 0.11 |
| **Induction therapy, %yes (n)** | 29 (136) | 25 (47) | 26 (19) | 0.66 |
| **PRA>50%, %yes (n)** | 6 (26) | 7 (12) | 17 (10) | 0.01 |
| **Flow crossmatch (%positive)** | 2.8 (13) | 4.9 (8) | 1.4 (1) | 0.34 |
| **Dual-organ (%yes)** | 8.8 (42) | 2.1 (4) | 2.7 (2) | <0.01 |

### Definition of rejection

In addition to scheduled surveillance EMBx, patients with clinically suspected rejection underwent for-cause EMBx, echocardiography, and serological testing. Acute cellular rejection (ACR) was defined by pathological grading of EMBx ≥ 2R per International Society of Heart and Lung Transplant (ISHLT) 2005 criteria.^30^ AMR was defined by at least two of the following: 1) immunohistochemical evidence of rejection on EMBx per ISHLT 2013 criteria (either positive C4d stain on immunofluorescence or presence of monocytes or CD68+ macrophages in the capillary endothelium), 2) presence of DSA (mean fluorescent intensity >1000 by solid phase assay) or antibodies targeting non-HLA antigens, and 3) symptoms of heart failure or graft dysfunction in absence of another etiology such as CAV.^10^ In addition, patients with isolated graft dysfunction and high clinical suspicion of AMR in the setting of ancillary data (e.g. hemodynamic compromise without another cause, cardiac MRI, dd-cfDNA, etc) were deemed to have AMR as well. In patients who were treated for AMR, we further stratified them by BP-AMR and BN-AMR. In many cases of AMR, a concurrent cellular component was observed. These patients with ‘mixed rejection’ were counted within the AMR cohort.^31^

### Rejection treatment and follow up

Confirmed or suspected episodes of rejection were treated at the discretion of the HT attending physician and individualized to each patient, although multidisciplinary meetings were held at both institutions weekly to discuss patients’ care. PLEX, when employed, consisted of five sessions, every other day. IVIG was given either as small doses (0.1-0.2mg/kg) between PLEX sessions or as larger boluses (1g/kg) at the end of PLEX, to achieve cumulative target dose 1.5-2g/kg.^28,29^ Rituximab was given as a single infusion at the end of treatment course (375mg/m^2^). Tocilizumab (8mg/kg) and bortezomib (1.3mg/m^2^) were variably given according to previously published studies.^26,27,29,32^ After treatment, patients underwent follow-up EMBx, echocardiography, and serological testing in 4-6 weeks. Additional interval testing was non-uniform and occurred at the discretion of the HT team.

### Outcomes

All patients were followed for at least one-year post-HT. The last date of censor was 9/1/2021 for Washington University in St. Louis and 10/1/2024 for Massachusetts General Hospital. The primary outcome was all-cause mortality. The secondary outcome was a composite death, re-transplantation, or new development of ISHLT grade 2 or 3 CAV as ascertained by coronary angiography.

### Statistical analysis

Categorical variables were reported as percent of total and compared using Chi-squared test or Fisher’s exact test as appropriate. Continuous variables were reported as mean (standard deviation) or median (interquartile range) and compared using t-test or Whitney-Mann test depending on the distribution. Kaplan-Meier curves were generated for no rejection, ACR-only, or AMR and compared via the log-rank test. In patients who experienced AMR, a Sankey diagram was created detailing their EMBx grade and clinical course (SankeyMATIC). In addition, we performed landmark analysis and generated Kaplan-Meier curves for the primary and secondary outcomes and compared BP-AMR and BN-AMR via the log-rank test. Finally, we performed a Cox proportional hazards analysis for death and the combination of death, retransplant, and ISHLT grade 2 or 3 CAV, adjusting for age, gender, race, presence of DSAs, and EMBx grading. Proportionality assumptions were tested using Schoenfeld residuals. All analyses were performed using R 4.4.2-4.5.0 and figures were created using GraphPad Prism (v10.0).

## Results

### Baseline characteristics of HT recipients

A total of 742 patients were included in this study. Of this cohort, 25% of patients had ACR-only and 10% of patients experienced at least one episode of AMR (**Table 1**). Compared to those without rejection or ACR, AMR patients were younger, more likely to be black, had higher prevalence of nonischemic cardiomyopathy, and greater degree of allosensitization. Prevalence of chronic kidney disease was highest in the ACR-only group and dual-organ recipients had very low rates of rejection.

In HT recipients who developed AMR, 33% of patients (N=24) did not have any evidence of rejection on EMBx by histology and immunohistochemistry. There were no significant baseline differences between BP-AMR and BN-AMR patients, although BN-AMR tended to occur later after HT (704 vs. 383 days, p=0.039, **Table 2**). Mixed rejection occurred in 22% (N=16) of patients.

**Table 2.**
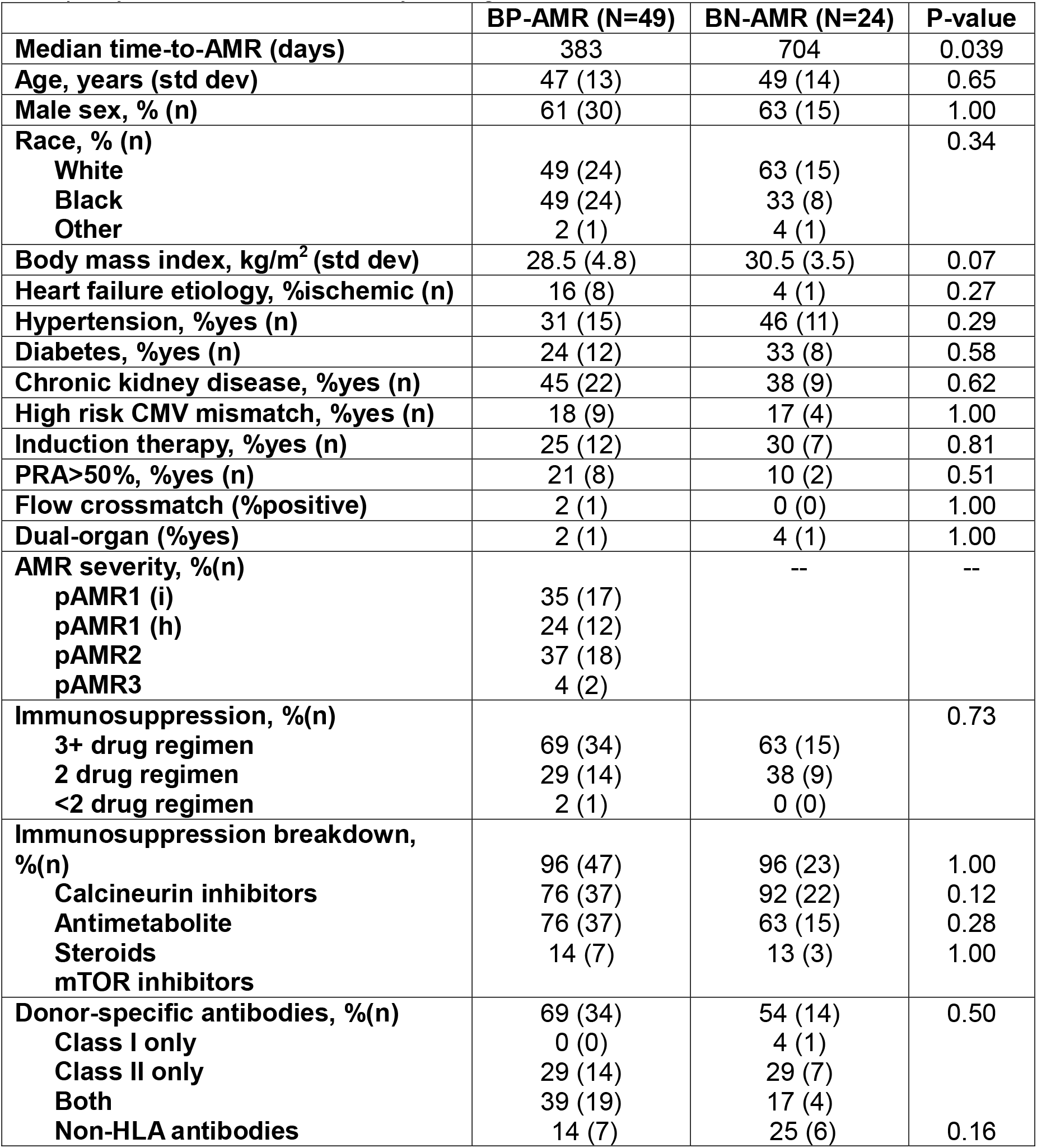
Baseline characteristics at time of antibody-mediated rejection (AMR), stratified by histopathological evidence on EMBx. High risk cytomegalovirus (CMV) mismatch: donor CMV IgG+ and recipient CMV IgG-, PRA: panel reactive antibodies, mTOR: mammalian target of rapamycin, HLA: human leukocyte antigens.

### AMR presentation

At baseline, BP and BN-AMR patients had normal graft function (mean LVEF 62%). At time of AMR diagnosis, 67% of patients had BP-AMR. Among this cohort, 60% of patients had pAMR1 and 37% had pAMR2 (**Table 2**). Most patients were taking at least 2-drug immunosuppression and there were no significant differences in the medication breakdown between BP-AMR and BN-AMR patients. The use of calcineurin inhibitors was high (96%) and mammalian target of rapamycin inhibitor was low (14%) in this population. Over 60% of patients had measurable DSAs with the detection of major histocompatibility (MHC) class II antibodies being the most common. There was no difference in the strength of the immunodominant DSA between the groups (**Supplemental table 2**) Sixteen percent of patients had non-HLA antibodies, with antibodies targeting angiotensin II type 1 receptor being the most common.

Compared to BP-AMR, BN-AMR patients had a significantly higher incidence of previously treated ACR (54% vs. 18%, p<0.01), although there were institutional differences (**Supplemental table 3**). BN-AMR patients had numerically lower left ventricular ejection fraction (LVEF, 42% vs. 48%, p=0.23), cardiac index (1.9 vs. 2.1 L/min/m^2^, p=0.21), and worse right ventricular function (p=0.19), although the differences were not statistically significant (**Figure 1A-C**). There was no difference in the central venous pressure, which was elevated in both cohorts consistent with clinical volume overload (15 vs. 15mmHg, p=0.45).

**Table 3.** Cox proportional hazards analysis of death (top) and death, retransplant, or International Society of Heart and Lung Transplant (ISHLT) grade 2 or 3 coronary artery vasculopathy (CAV). Presence of immunohistological evidence of antibody-mediated rejection was not associated with death but was associated with the composite outcome of death, retransplant, and ISHLT grade 2 or 3 CAV.

|  | **Hazard ratio** | **Confidence interval** | **P-value** |
| --- | --- | --- | --- |
| **Death** |  |  |  |
| Age | 1.01 | 0.97-1.04 | 0.74 |
| Male sex | 1.44 | 0.62-3.32 | 0.39 |
| White race | 0.96 | 0.43-2.11 | 0.91 |
| Donor-specific antibody | 0.88 | 0.35-2.24 | 0.79 |
| Positive biopsy | 2.04 | 0.72-5.75 | 0.18 |
| **Death, retransplant, ISHLT grade 2 or 3 CAV** |  |  |  |
| Age | 1.00 | 0.98-1.03 | 0.77 |
| Male sex | 1.37 | 0.66-2.82 | 0.40 |
| White race | 1.10 | 0.54-2.22 | 0.80 |
| Donor-specific antibody | 0.90 | 0.41-2.00 | 0.80 |
| Positive biopsy | 2.76 | 1.09-7.00 | 0.03 |

**Figure 1.**
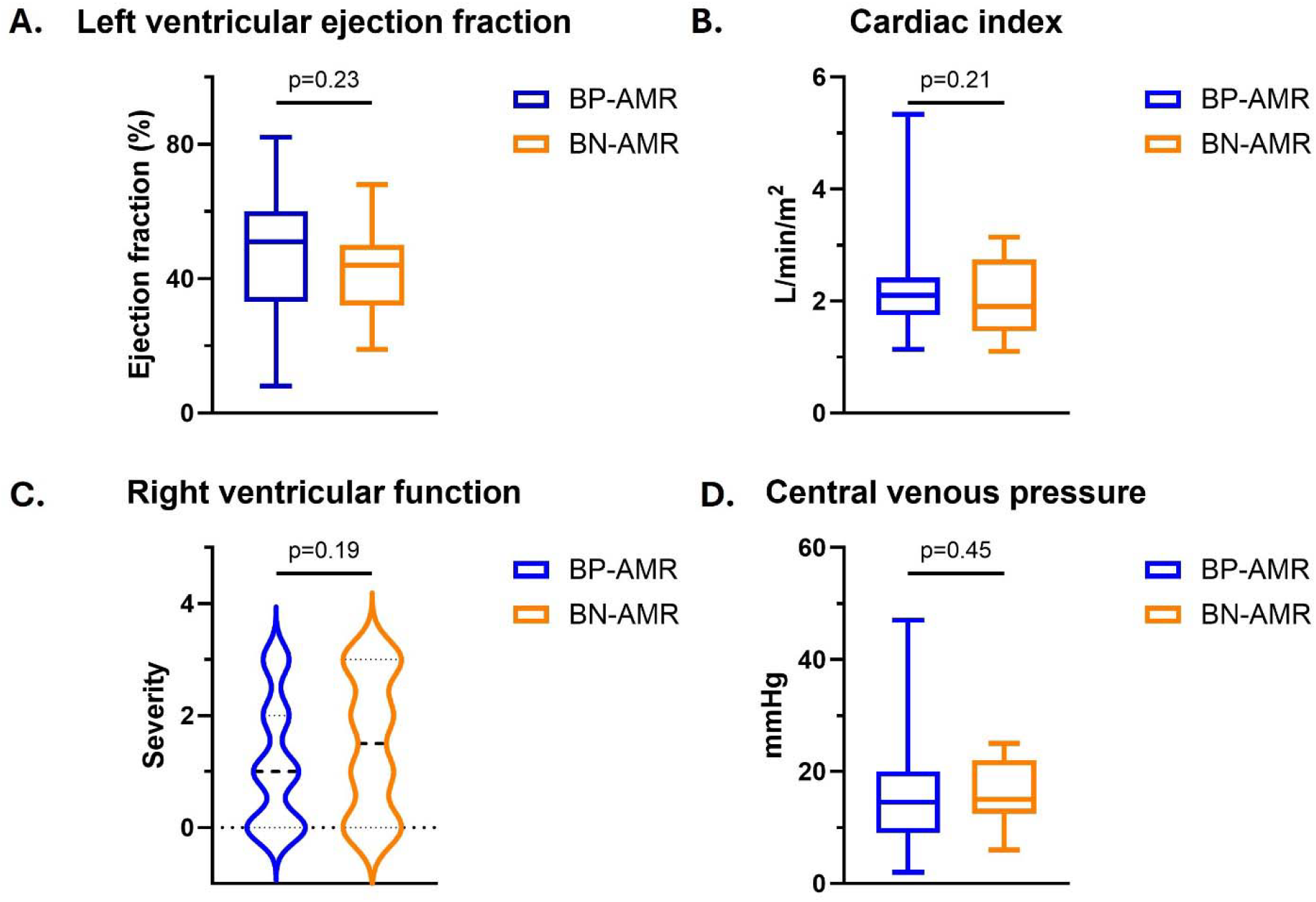
Echocardiographic and hemodynamic parameters at time of antibody-mediated rejection (AMR) diagnosis. Patients with BN-AMR had numerically lower left ventricular ejection fraction, cardiac index, and worse right ventricular function compared to BP-AMR (**panels A-C**). BN-AMR and BP-AMR patients had similarly elevated central venous pressures (**panel D**). Right ventricular function was defined semi-quantitatively on transthoracic echocardiogram as normal (0), mild (1), moderate (2), or severe (3) dysfunction by echocardiography-boarded physicians.

### AMR treatment and clinical course

After diagnosis, patients were treated with various modalities for their AMR (**Supplemental table 4**). Intravenous steroids were the most common therapy, followed by PLEX and IVIG. Rituximab was given in 70% of cases. Compared to BP-AMR, patients with BN-AMR received numerically less steroid pulse (79% vs. 90%), PLEX (75% vs. 90%), and IVIG (79% vs. 94%, **Figure 2a**). Total use of biologics such as rituximab and tocilizumab was similar. More specifically, different combinations were employed and tailored to each patients’ circumstances, although the most common regimen of PLEX, IVIG, and rituximab was employed in over 60% of patients.

**Figure 2.**
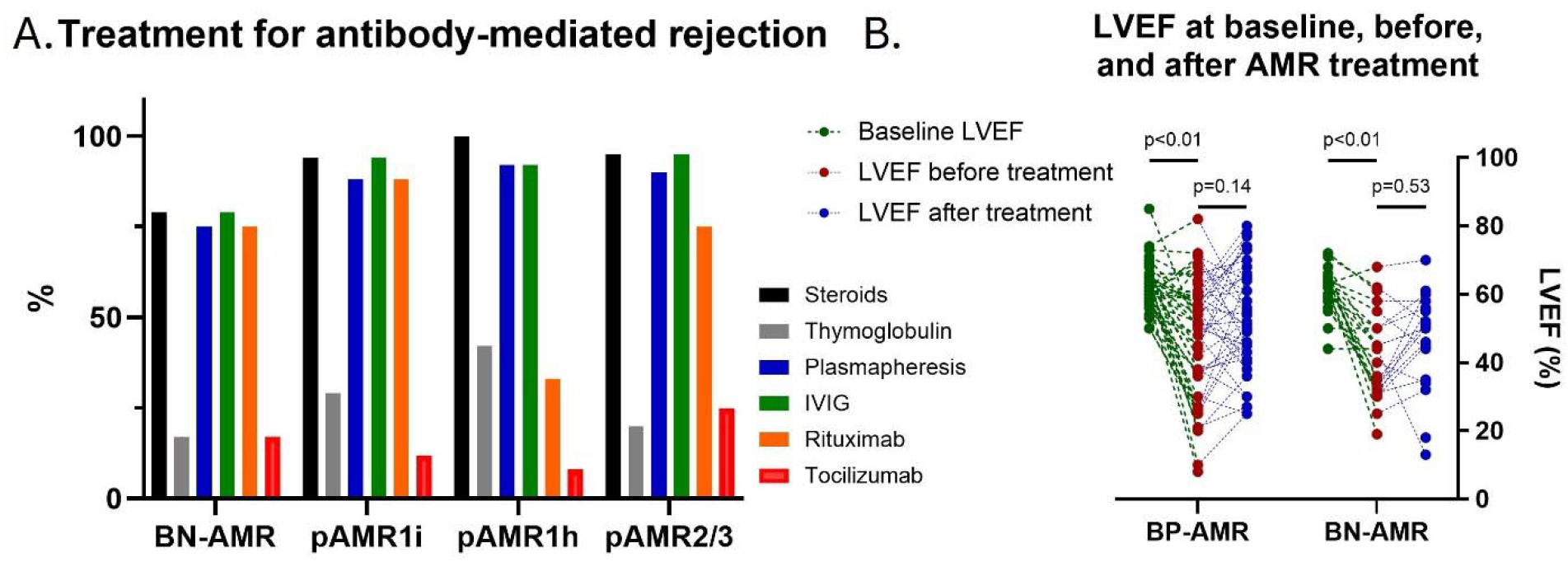
Treatment and left ventricular ejection fraction (LVEF) response for BP-AMR and BN-AMR patients. AMR patients were treated with various modalities, most commonly a combination of steroids, plasmapheresis, intravenous immunoglobulin (IVIG), and rituximab. BN-AMR patients received numerically less steroids, plasmapheresis, and IVIG (**panel A**). The use of thymoglobulin, rituximab, and tocilizumab were similar between BP and BN-AMR. There was no difference in baseline graft function (62% vs. 62%, p=0.94) and both BP and BN-AMR patients experienced a significant reduction in LVEF from baseline to AMR diagnosis (p<0.01, **panel B**). After treatment, BP-AMR patients experienced a trend towards increase in LVEF (48% to 53%, p=0.14) whereas BN-AMR patients had essentially unchanged LVEF (A, 42% to 46%, p=0.536).

After treatment, patients with BP-AMR experienced a nonsignificant trend towards LVEF improvement (48% to 53%, p=0.14) whereas patients with BN-AMR had essentially unchanged graft function (LVEF 42% vs. 46%, p=0.53, **Figure 2b**). There was significant heterogeneity in the response to treatment between groups. Despite aggressive treatment, morbidity and mortality were high. Over 20% of patients died during index hospitalization. In those who survived to hospital discharge, 40% of patients continued to have HF symptoms or graft dysfunction at follow up (**Figure 3**).

**Figure 3.**
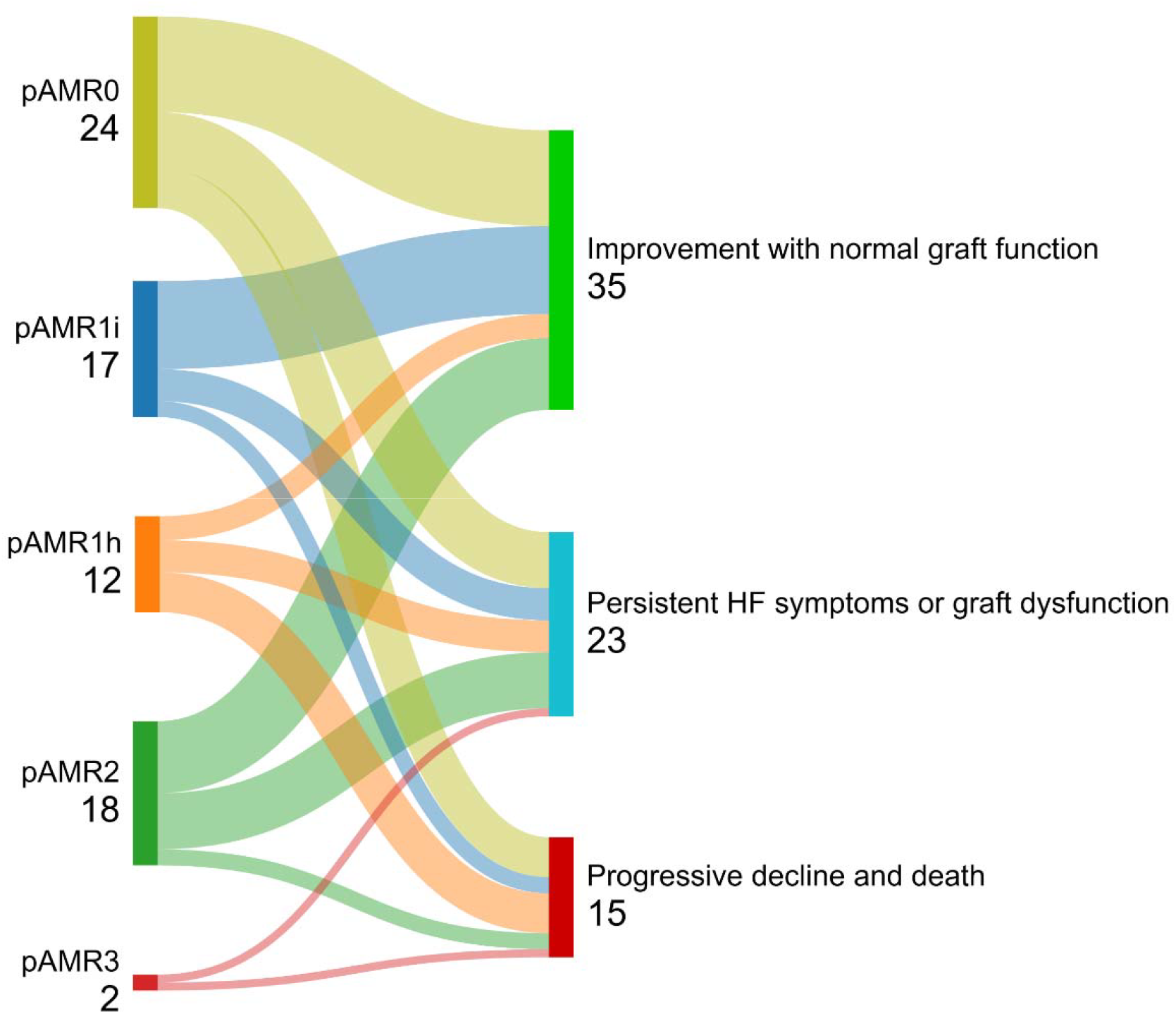
Clinical course of antibody-mediated rejection stratified by biopsy grading. Figure created with SankeyMATIC.

### Long-term clinical outcomes

In landmark analysis, the conditional 5-year survival in the overall cohort was above 80% at both institutions (**Supplemental figure 1**). Patients who experienced AMR had significantly higher mortality whereas patients who experienced ACR-only had similar survival to the no rejection group (**Figure 4**). In Kaplan-Meier analyses, there was no difference in all-cause mortality between BP and BN-AMR, with median 5-year survival after AMR treatment ranging between 50-60% (**Figure 5a)**. The majority of death events were due to cardiac causes (**Supplemental table 5**). There was no difference in the composite outcome of death, retransplant, or new development of ISHLT grade 2 or 3 CAV either (**Figure 5b**). Finally, in Cox proportional hazards analysis, BP-AMR was not associated with death (HR 2.04, CI 0.72-5.75, p=0.18) although was associated with the combined outcome of death, retransplant, or ISHLT grade 2 or 3 CAV (HR 2.76, CI 1.09-7.00, p=0.03). Age, sex, race, and DSAs were not associated with either outcome (**Table 3**).

**Figure 4.**
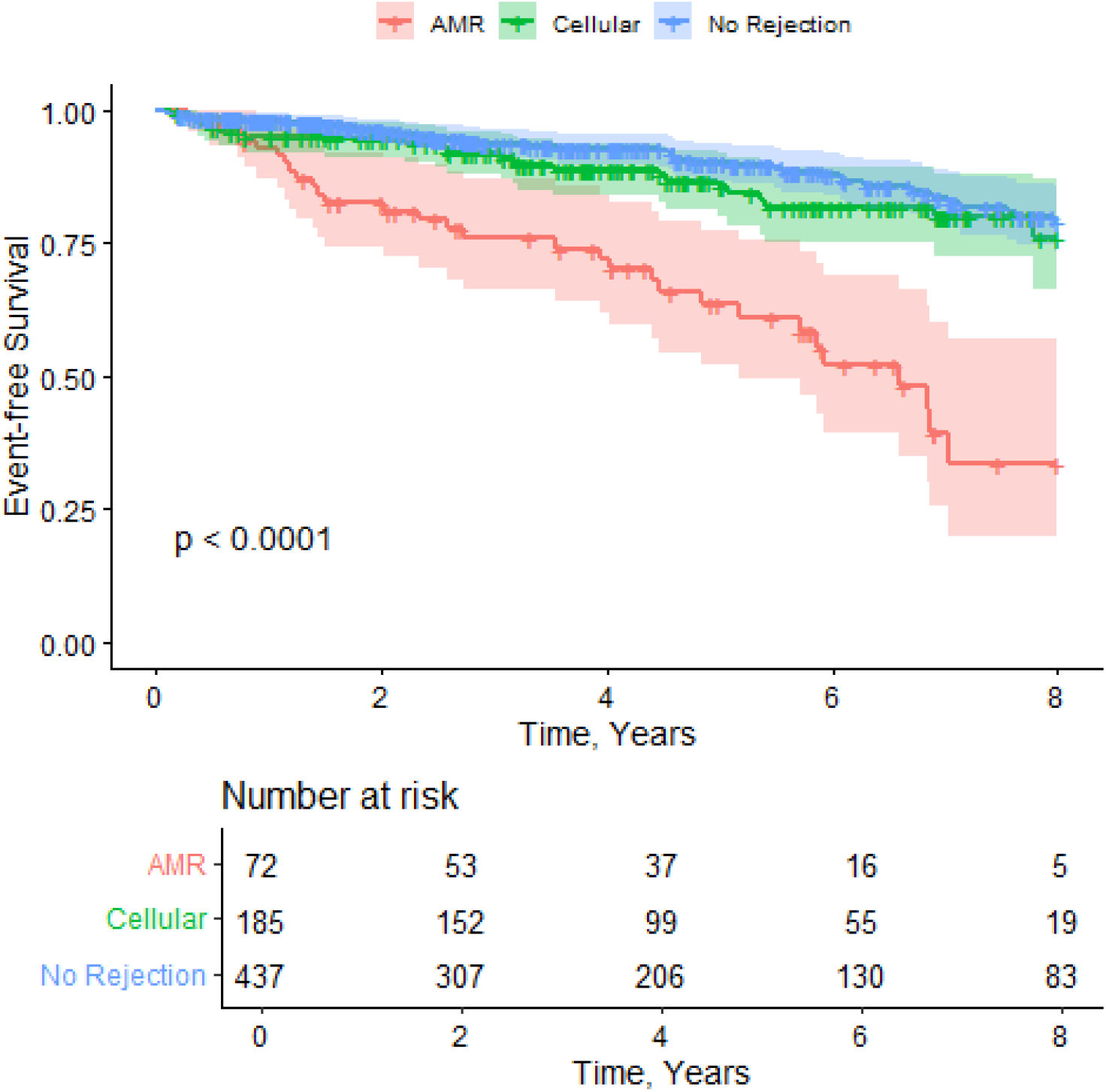
All-cause mortality stratified by rejection. Patients with cellular rejection had equal survival to those without rejection. Patients with antibody-mediated rejection had significantly worse survival (log rank p<0.01).

**Figure 5.**
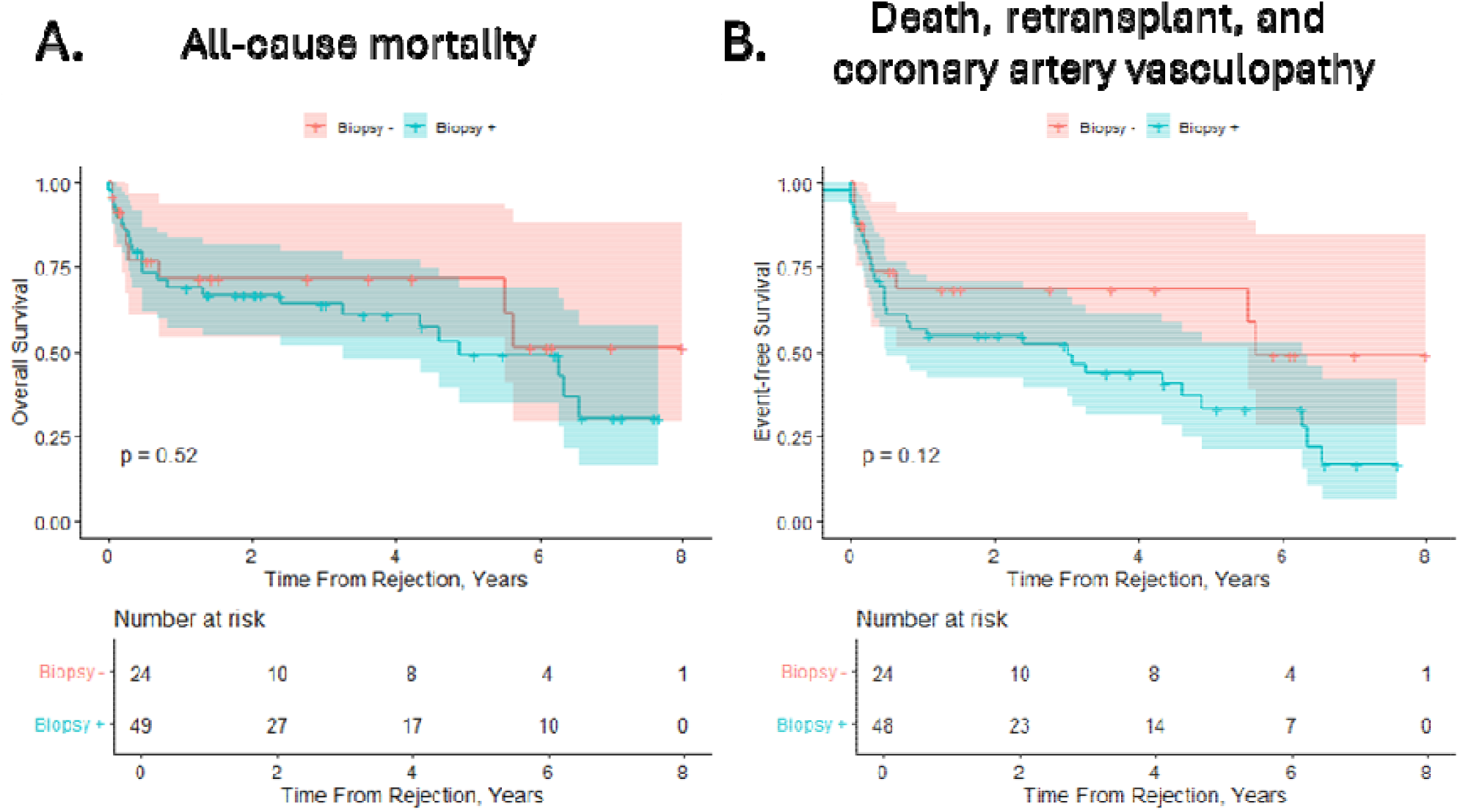
All-cause mortality and composite of death, retransplant, and International Society of Heart and Lung Transplant (ISHLT) grade 2 or 3 coronary artery vasculopathy (CAV) after treatment of antibody-mediated rejection (AMR). There was no difference in all-cause mortality between BP and BN-AMR (**panel A**, log rank p=0.66). BP-AMR patients showed a trend towards a higher rate of the composite outcome (**panel B**, log rank p=0.12).

## Discussion

In this contemporary, 2-center study of almost 750 *de novo* HT recipients, we describe in detail the clinical presentation, treatment, and outcomes of patients with AMR. We found that AMR occurs late after transplant and is associated with decreased survival compared to those with ACR or no rejection. Over one third of patients with AMR had no histopathological evidence of rejection on EMBx. These BN-AMR episodes were diagnosed even later, treated less aggressively, and had equally poor long-term outcomes compared to ‘classic’ BP-AMR.

First described in 1990s as “biopsy-negative” or “humoral” rejection, the pathologic and clinical definitions of AMR have evolved over time.^8,10^ Previous studies in this space were mostly single-centered and focused on patients who had a positive EMBx.^18,33^ Few reports have evaluated the treatment and clinical course of patients with high suspicion for AMR but without histological evidence of rejection (BN-AMR). Our study adds to the existing literature by demonstrating a ‘negative’ biopsy does not preclude the diagnosis of AMR and BN-AMR is common in a real-world setting. In fact, we found that BN-AMR patients (judged not to have another etiology responsible for their diagnosis by the HT team) had in-hospital mortality or persistent graft dysfunction of 50%. More importantly, BN-AMR was less aggressively treated, likely related to the diagnostic uncertainty, and in Kaplan-Meier and Cox proportional hazards analyses, had similar risk of mortality as BP-AMR at long-term follow up. As such, a high index of suspicion is necessary to prevent the high morbidity and mortality associated with AMR, especially for those with BN-AMR.

We also found that BN-AMR presented at later timepoints following HT than BP-AMR. This could signify that these patients were more advanced in their disease or had a more chronic inflammatory process. In addition, it could be that BN-AMR was the result of prior rejection injury that persisted or resolved, a concept that is partly supported by the fact that BN-AMR patients were much more likely to have been treated for a previous episode of ACR. Although LVEF was similarly preserved in BP and BN-AMR patients before their AMR episode and severe CAV was not present, BN-AMR patients may have accrued subclinical microvascular disease or early CAV that went unrecognized. These could in part contribute to the non-statistically significant lower LVEF, cardiac index, worse RV function, and more modest LVEF improvement in this group even after treatment. We noted that BP-AMR was associated with the composite outcome of death, retransplant, and ISHLT grade 2 or 3 CAV in our Cox proportional hazard analysis, driven in large part by CAV. Although this should be interpreted with caution, we hypothesize that this may be due to the earlier presentation and increased surveillance, especially of CAV, after AMR treatment. Interestingly, we found that although DSAs were similar between BP-AMR and BN-AMR (69% vs. 54%, p=0.50), BP-AMR patients had numerically higher incidence of MHC class II DSAs (67% vs. 47%). This may also contribute to the trend of our composite outcome, given the known association between class II DSAs and CAV.

Overall, the prevalence of AMR in this study was lower than some previous reports, although similar to another multicentered study looking at CAV trajectories.^34^ We suspect this is due to under-recognition of subclinical AMR. Due to the timeframe of this study, most patients were treated for AMR in an era before dd-cfDNA and Molecular Microscope Diagnostic System (ThermoFisher, MMDx) were widely available. These novel tools could allow for earlier detection of AMR in HT recipients. Moreover, the combination of tissue transcriptomic analysis and dd-cfDNA may be more sensitive at identifying AMR than traditional EMBx and histological examination.^23,35-37^ This is in line with a study by Goldberg et al showing that dd-cfDNA can be used to identify BN-AMR in the Genomic Research Alliance for Transplantation Study (GRAfT) participants, which occurred in 38x% of treated AMR patients.^38^ Our study complements the GRAfT consortium findings but further describes the various treatments received and short and long-term outcomes in a larger population. More recently, artificial intelligence-based imaging analysis have also been shown to increase the diagnostic yield in identifying AMR.^39^ Ultimately, incorporating clinical, tissue-specific (EMBx/MMDx), and serological information (DSA/dd-cfDNA) in a multimodal fashion may allow for greater insight into pre/subclinical stages of AMR and lead to earlier diagnosis and treatment.^40^

Finally, despite aggressive treatment, long-term prognosis after AMR is poor, with 5-year mortality approaching 50% after diagnosis in our cohort. We found that the treatment protocols for individual patients and response to treatment were highly variable. This lack of uniformity in AMR management nevertheless reflects real-world practices and future research should focus on phenotyping AMR and investigating whether specific treatments may benefit a particular phenotype. While we found that BP-AMR patients experienced a more pronounced improvement in LVEF, another area of future research will be to define the clinical and functional variables that may reflect response to treatment and correlate with long-term outcomes.

This study has several limitations. First, our results may not be generalizable to other institutions, given differences in the protocols for immunosuppression, surveillance, and management which could all impact AMR diagnosis and post-treatment outcomes. However, this heterogeneity reflects current clinical practice and reporting a real-world experience in a large sample size is a strength of our study. Second, it is possible some BN-AMR patients did not actually have AMR and rather nonspecific graft dysfunction. We acknowledge that this distinction is difficult to prove and there may be clinical overlap. However, BN-AMR patients had similarly poor clinical outcomes as BP-AMR and in exploratory analysis of BN-AMR patients who had dd-cfDNA data available (N=5), the mean dd-cfDNA was 2.1%. Thus, we favor that our BN-AMR patients actually had true rejection, rather than nonspecific graft dysfunction. Importantly, routine use of dd-cfDNA in conjunction with MMDx may help identify patients with BN-AMR who do not have rejection and thus prevent unnecessary treatment. Third, cardiac MRI was not routinely performed at our centers. Advanced imaging may be useful to identify rejection in BN-AMR patients and should be further studied in this population.^41,42^ Finally, our study was not powered to make conclusions about efficacy of various AMR treatments. Yet, this is one of the largest AMR reports to date over an extended period of time and detailing patients’ clinical course and outcomes will be of interest to the HT community. Future studies should focus on identifying whether specific treatments may improve outcomes in different phenotypes of AMR.

In summary, AMR after HT is associated with poor clinical outcomes. BN-AMR is common and a high index of suspicion is necessary for diagnosis and prompt treatment. Future studies should focus on early detection, delineating the pathophysiology and phenotypes of AMR, and investigating the optimal mode of treatment.

## Data Availability

All data produced in the present study are available upon reasonable request to the authors.

## Abbreviations

AMR: antibody-mediated rejection
HT: heart transplant
EMBx: endomyocardial biopsy
DSAs: donor-specific antibodies
CAV: coronary artery vasculopathy
dd-cfDNA: donor-derived, cell-free DNA
PLEX: plasmapheresis/exchange
IVIG: intravenous immunoglobulin
BP-AMR: biopsy-positive AMR
BN-AMR: biopsy-negative AMR
ACR: acute cellular rejection
ISHLT: International Society of Heart and Lung Transplant
MHC: major histocompatibility complex
LVEF: left ventricular ejection fraction
GRAfT: Genomic Research Alliance for Transplantation Study

## Acknowledgement

The authors would like to thank the Mass General Brigham Spark Award for funding this study.

