## Supplemental material for "Clinical course and outcomes of antibody-mediated rejection after heart transplant in the contemporary era"

**Supplemental materials**

**Supplemental table 1. Surveillance protocols at both institutions.** X denotes routine surveillance endomyocardial biopsies.

| **Weeks after transplant** | **Barnes-Jewish Hospital/**  **Washington University in St. Louis** | **Massachusetts General Hospital/**  **Harvard Medical School** |
| --- | --- | --- |
| **1** | **x** | **x** |
| **2** | **x** | **x** |
| **3** | **x** |  |
| **4** |  | **x** |
| **6** | **x** |  |
| **8** | **x** | **x** |
| **10** | **x** |  |
| **12** | **x** |  |
| **16** | **x** | **x** |
| **20** | **x** |  |
| **24** |  |  |
| **28** | **x** | **x** |
| **52** | **x** | **x** |
| **78** |  | **x** |
| **104** |  | **x** |
| **156** |  | **x** |
| **208** |  | **x** |
| **260** |  | **x** |

**Supplemental table 2. Strength of immunodominant donor-specific antibody (DSA), using 5000 mean fluorescent intensity (MFI) as cutoff.** There was no difference in the strength of the immunodominant DSAs between BP and BN-AMR (p=1.00). Expressed as percentage of patients who had DSAs.

| **Strength of DSAs, %(n)** | **BP-AMR** | **BN-AMR** |
| --- | --- | --- |
| **< 5000 MFI** | **26 (9)** | **21 (3)** |
| **≥ 5000 MFI** | **74 (26)** | **79 (11)** |

**Supplemental table 3. Previously treated episodes of cellular rejection, stratified by institution.** BP-AMR patients were less likely to be treated for a previous episode of cellular rejection (18% vs. 54%, p<0.01). There was a significant institutional difference, with patients at Washington University in St. Louis (WashU) having much lower incidence of treated cellular rejection than those at Massachusetts General Hospital (MGH).

|  | **BP-AMR** | | | **BN-AMR** | | |
| --- | --- | --- | --- | --- | --- | --- |
|  | WashU | MGH | P-value | WashU | MGH | P-value |
| Previous cellular rejection, %(n) | 5 (2) | 35 (7) | <0.01 | 38 (6) | 88 (7) | 0.02 |

**Supplemental table 4. Treatments received for BP- and BN-AMR patients.** Patients with BN-AMR received numerically less steroids (79% vs. 90%), plasmapheresis (75% vs. 90%), and IVIG (79% vs. 94%). The use of thymoglobulin, rituximab, and tocilizumab were similar.

|  | **BP-AMR (N=49)** | **BN-AMR (N=24)** | **P-value** |
| --- | --- | --- | --- |
| **Treatment, %(n)**  **Pulse steroids**  **Thymoglobulin**  **Plasmapheresis/exchange**  **Intravenous immunoglobulin**  **Rituximab**  **Tocilizumab**  **Other** | 90 (44)  24 (12)  90 (44)  94 (46)  69 (34)  17 (8)  2 (1) | 79 (19)  21 (5)  75 (18)  79 (19)  75 (18)  17 (4)  0 (0) | 0.28  1.00  0.16  0.11  0.78  1.00  1.00 |

**Supplemental table 5. Cause of death in patients with BP and BN-AMR.** Most death events occurred due to cardiac causes and there was no statistical difference between BP and BN-AMR (p=0.57). Expressed as percentage of total number of patients in each cohort.

| **Cause of death, %(n)** | **BP-AMR** | **BN-AMR** |
| --- | --- | --- |
| **Cardiac** | **33 (16)** | **30 (7)** |
| **Malignancy** | **0 (0)** | **0 (0)** |
| **Infection** | **8 (4)** | **0 (0)** |
| **Other** | **12 (6)** | **4 (1)** |

**Supplemental figure 1. Conditional 5-year survival, stratified by institution.** MGH, Massachusetts General Hospital, WUSTL, Washington University in St. Louis.

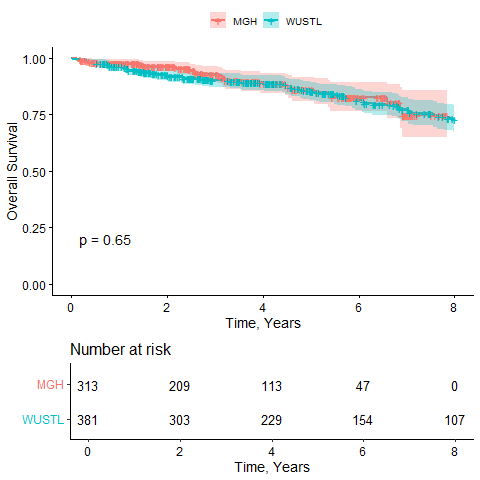
